# Lower limb muscle activity underlying temporal gait asymmetry post-stroke

**DOI:** 10.1101/19010421

**Authors:** Gabriela M. Rozanski, Andrew H. Huntley, Lucas D. Crosby, Alison Schinkel-Ivy, Avril Mansfield, Kara K. Patterson

**Author notes:** **Corresponding author** Gabriela M. Rozanski, Toronto Rehabilitation Institute, 550 University Avenue, 11-107, Toronto, Ontario M5G 2A2, Canada. **Declarations of interest** None.

## Abstract

**Objective:** Asymmetric walking after stroke is common, detrimental, and difficult to treat, but current knowledge of underlying physiological mechanisms is limited. This study investigated electromyographic (EMG) features of temporal gait asymmetry (TGA).

**Methods:** Participants post-stroke with or without TGA and control adults (n=27, 8, and 9, respectively) performed self-paced overground gait trials. EMG, force plate, and motion capture data were collected. Lower limb muscle activity was compared across groups and sides (more/less affected). Correlation between burst timing variables and asymmetry ratios was examined.

**Results:** Significant group by side interaction effects were found: fewer TGA group dorsiflexor bursts during swing (p=.0009), more affected plantarflexor stance activity ended early (p=.0006) and less affected dorsiflexor on/off time was delayed (p<.01) in persons with asymmetry compared to symmetric and normative controls. Less affected side EMG timing correlated most with swing time ratio (r=0.68-0.90, p<.001).

**Conclusions:** Temporal patterns of muscular activation, particularly about the ankle around the stance-to-swing transition period, are associated with TGA. The results may reflect specific impairments or compensations that affect locomotor coordination.

**Significance:** Neuromuscular underpinnings of spatiotemporal asymmetry have not been previously characterized. These novel findings may inform targeted therapeutic strategies to improve gait quality after stroke.

## 1. Introduction

Functional mobility is often affected after stroke due to impairments in walking, which can restrict activity and participation (Andrenelli et al. 2015). Therefore, improving hemiparetic gait is a major focus of rehabilitation (Jette et al. 2005; Latham et al. 2005) and patients’ top goal (Bohannon et al. 1988). While speed and endurance may increase with physical therapy, ambulatory dysfunction often persists; more than half of community-dwelling individuals with chronic stroke walk asymmetrically (Patterson et al. 2008a). This has clinical implications; a number of long-term negative consequences have been proposed (Patterson et al. 2008a), including musculoskeletal pathology (Jorgensen et al. 2000; Norvell et al. 2005; Beaupre and Lew 2006), falls due to instability (Lewek et al. 2014; Lien et al. 2017), and inefficient energy consumption (Ellis et al. 2013; Awad et al. 2015) leading to reduced activity (Michael et al. 2005). Furthermore, walking appearance is important to people with stroke (Bohannon et al. 1991). Unfortunately, asymmetric gait can be resistant to intervention (Patterson et al. 2015) and may even worsen over time (Turnbull and Wall 1995; Patterson et al. 2010a).

Spatiotemporal parameters are used to describe the quality of hemiparetic gait and may indicate specific biomechanical deficits or compensations (Allen et al. 2011) with respect to kinetics (Kim and Eng 2003), coordination (Wonsetler and Bowden 2017), and muscle function (Balasubramanian et al. 2007). Temporal gait asymmetry (TGA), measured as a ratio of the more and less affected side swing (or stance) time, is related to paretic ankle muscle strength and spasticity (Hsu et al. 2003; Lin et al. 2006) as well as lower limb impairment due to stroke (Brandstater et al. 1983; Patterson et al. 2008b); however, these findings are limited by the use of gross clinical assessments or conditions that are not fully reflective of complex gait dynamics. Good motor recovery or fast walking speed does not necessarily imply symmetry and age is not a factor (Patterson et al. 2008b, 2012). More ecologically valid and direct measurements are necessary to reveal the underlying neural mechanisms or adaptation strategies that result in TGA. With better understanding of this detrimental outcome of stroke, novel targeted therapy can be developed to improve symmetry and advance the field of gait rehabilitation.

Surface electromyography (EMG) has long been used to study neuromuscular control of locomotion after stroke. Recorded muscle activity, representing nervous system output, can be quantified in various ways and compared with healthy controls or the less-affected side in the context of hemiparesis to elucidate the resulting aberrant motor behaviour. Despite considerable inter-subject variability (Knutsson and Richards 1979), some common impairments have been identified (Olney and Richards 1996): absence or reduced amplitude of tibialis anterior activation, prolonged quadriceps and hamstring firing, and premature calf muscle onset. Lower limb agonist/antagonist co-contraction and synergistic coordination during hemiparetic walking have also been examined (Rosa et al. 2014). Yet, the association of these patterns with gait symmetry has not been directly investigated. It is still unclear how neurophysiological changes due to stroke affect the quality of gait. This study aimed to determine whether lower limb muscle activity and timing (i.e. burst on/offset) in persons with TGA is different than symmetric post-stroke individuals and healthy adults. The results could elucidate specific EMG patterns during overground walking that underlie temporal asymmetry as well as inform future clinical approaches to gait assessment and treatment.

## 2. Methods

### 2.1 Design and subjects

This study was a cross-sectional analysis of data from two research trials approved by the University Health Network Research Ethics Board (study IDs 14-7428 and 15-9523). Both examined community-dwelling individuals with stroke. The first was a randomized controlled trial of perturbation training (registration #: ISRCTN05434601) (Mansfield et al. 2015) from April 2014 to August 2017. Of the 90 enrolled participants, 20 completed a gait assessment (see below). The second, an ongoing observational study of gait and rhythm ability that began July 2016, also included healthy adult volunteers (n=10 control subjects plus 29 individuals with stroke at the time of this publication). All participants in the current study were able to walk 10 m without a gait aid or physical assistance. Relevant exclusion criteria were: pre-existing or other neurological or musculoskeletal condition(s) significantly impacting gait (e.g. osteoarthritis, Parkinson’s disease); cognitive, language or communication impairment; and recent major illness, injury, or surgery. Written informed consent was obtained from all participants prior to data collection.

### 2.2 Data collection and assessment procedures

#### 2.2.1 Demographics and clinical assessment

The following information, as applicable, was recorded at the time of enrolment: age, sex, time post-stroke, stroke location, and pre-morbid medical history. Participants with stroke also underwent a battery of standardized clinical tests administered by a trained research assistant. Relevant to this study, the Chedoke McMaster Stroke Assessment (CMSA) was used to measure motor impairment of the lower extremity; this tool has good reliability and concurrent validity (Gowland et al. 1993).

#### 2.2.2 Overground gait

Detailed gait assessment was conducted at Toronto Rehabilitation Institute in FallsLab, a 6 m x 3 m movable platform (static for the duration of data collection) equipped with passive motion capture (Vicon Motion Systems Ltd, Oxford, UK). Participants were full-body outfitted with reflective markers on anatomical landmarks (only one was used for kinematic analysis, see 2.3.1). An eight-channel wireless EMG system (Noraxon USA Inc., Scottsdale, Arizona, USA) measured activity from four pairs of lower limb muscles. Electrodes were placed bilaterally on clean skin (abraded and swabbed with rubbing alcohol) according to SENIAM recommendations (Hermens et al. 2000) and Criswell locations (Cram and Kasman 2011) for tibialis anterior (TA, lateral to the tibia approximately 1/3 of the distance between the ankle and knee), medial gastrocnemius (MG, 2 cm medial to the midline just distal from the knee), rectus femoris (RF, middle of the thigh approximately halfway between the knee and anterior superior iliac spine), and biceps femoris (BF, 2 cm lateral to the midline approximately halfway between the gluteal fold and posterior knee). Participants walked at a self-selected pace across a platform comprised of eight (two by four) 1.5 m x 1.5 m force plates (Advanced Mechanical Technology Inc., Watertown, Massachusetts, USA) while wearing an integrated active safety harness that allowed free movement without providing body weight support. Only the middle two pairs of force plates (four total) actively recorded, but gait was initiated before and terminated after (approximately 1 m) to allow for acceleration and deceleration. Five or six trials (depending on study protocol) were performed from the same marked starting point to position participants such that the right foot landed on the right side of the platform and vice versa. Rest breaks were given as needed. Force plate and electromyographic data were sampled at 250 Hz and 1000 Hz, respectively, synchronously with a custom Simulink (The Mathworks, Natick, Massachusetts, USA) program, and stored for offline processing. Motion files were captured separately at 100 Hz using the manufacturer’s Nexus software (Vicon Motion Systems Ltd, Oxford, UK).

### 2.3 Data processing and analysis

#### 2.3.1 Gait parameters

Force plate and motion capture data (labelled in Vicon Nexus v.1.8.5) were processed with Visual 3D v.5 (C-Motion Inc., Germantown, Maryland, USA). After applying a zero-lag 4^th^-order Butterworth low-pass filter (cutoff frequency: 25 Hz) (Winter 1990; Kram et al. 1998), vertical ground reaction forces were analyzed to determine foot-on and -off times using a 20 N threshold, then confirmed via visual inspection. Gait events for all but the first trial (to account for familiarization to the assessment procedure) (Boudarham et al. 2013) were extracted following visual inspection. One stride or gait cycle was between consecutive foot-on events and stance was the phase from foot-on to foot-off (expressed in absolute duration and as a percent of stride). Left and right swing times were computed as the duration between foot-off and the subsequent foot-on, then averaged over all strides. The swing time ratio (larger of the left and right side mean values in the numerator) served as the measure of TGA (Patterson et al. 2010b). Gait velocity was derived from the anterior motion of one pelvis marker (interpolated then low-pass Butterworth filtered at 6 Hz) (Yang and Pai 2014) and the average global magnitude was calculated for each processed trial.

#### 2.3.2 Electromyography

Custom programs written in MATLAB (R2015b; The Mathworks, Natick, Massachusetts, USA) were used to process raw EMG recordings after visual inspection for data quality. A zero-lag 4^th^-order bandpass Butterworth filter with cutoff frequencies of 10 Hz and 300 Hz was applied, followed by detrending, rectification, and low-pass filtering at 30 Hz (zero-lag 4^th^-order Butterworth) (Merletti 1999). The processed signal was scanned for EMG bursts, as defined by amplitude above the trial mean for at least 30 ms. Bursts that were separated by less than 30 ms were merged, as shorter muscle activations are generally considered to have no biomechanical effect (Bogey et al. 1992). These threshold parameters identified periods of activity with a balance of specificity and sensitivity such that the output appeared to accurately represent the raw data (Figure 1). Timing of the beginning (ON) and end (OFF) of the bursts from each muscle were extracted after visual inspection of every stride, and then expressed as a percentage of stride relative to the respective foot-on event (i.e., start of stance phase). Normative muscle activity patterns were identified from the healthy adult control data and allowed classification of bursts with respect to the phase of gait (i.e., stance and swing) as described below. A burst that began during stance and ended in the swing phase was considered part of the stance-to-swing transition period.

**Figure 1.**
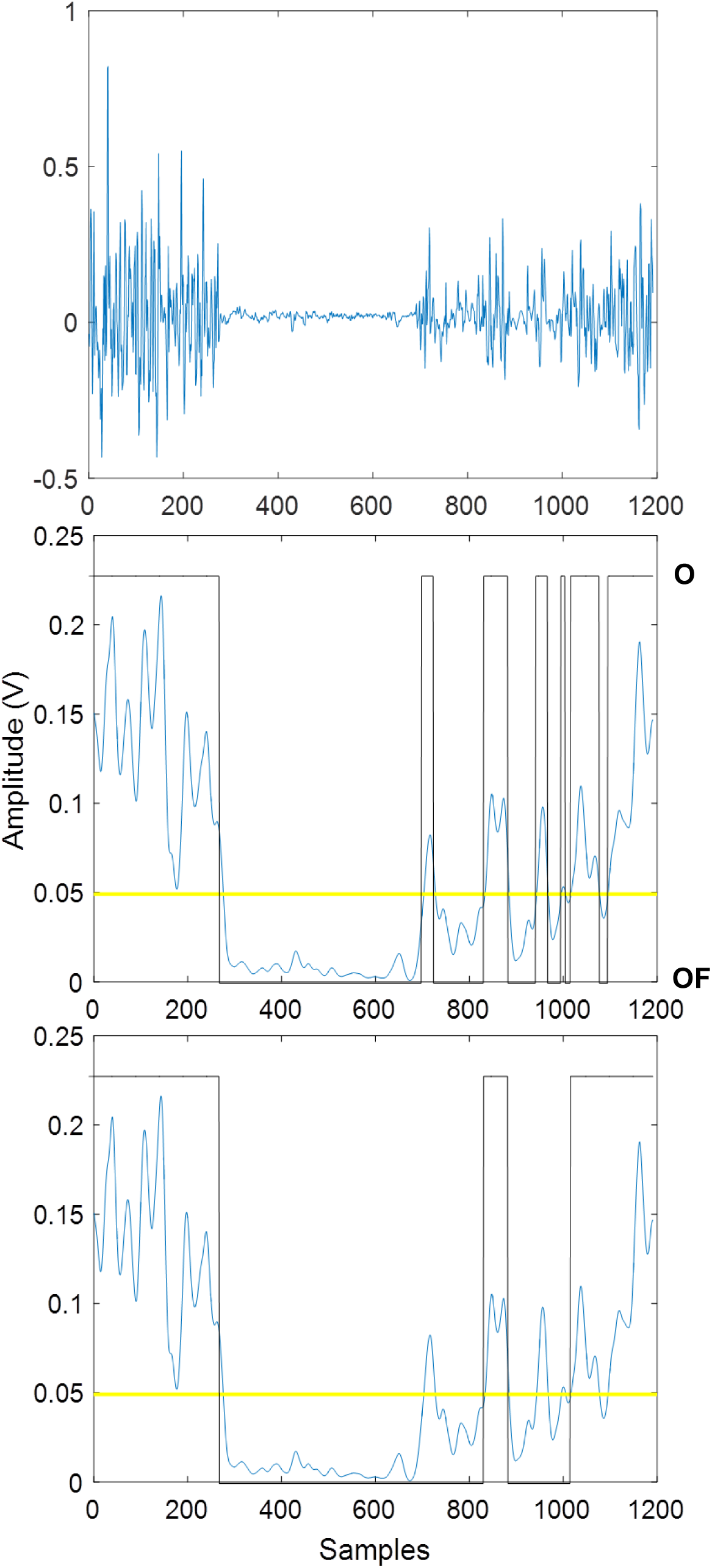
Electromyographic data processing. Raw EMG recording of a complete gait cycle sampled at 1000 Hz (top) was filtered, detrended and rectified. Conditioned signal (middle, blue) was scanned for muscle bursts (black) with amplitude above the threshold (trial mean, yellow line). Post-processing rejected any transitions that are less than 30 ms (bottom).

### 2.4 Statistical analysis

Participants with stroke were classified as either symmetric or asymmetric in swing time ratio based on the normative threshold of 1.06 (Patterson et al. 2010b). The more and less affected sides were designated according to self-report and clinical presentation. For the control adults, limbs were differentiated by swing time (higher = ‘more affected’, lower = ‘less affected’). Consequently, there was a between-subject factor of group (three levels: asymmetric, symmetric, and controls) and a within-subject factor of side with two levels. SAS 9.4 (SAS Institute, Cary, North Carolina, USA) software was used to describe the data (mean and standard deviation for continuous measures; frequency and percentages for categorical variables) and to perform statistical tests. Burst count outcomes were analyzed using negative binomial regression with group, side, and group*side as predictors. A repeated measures ANOVA was performed on the gait parameters and burst timing continuous variables that approximated a normal distribution after visual examination. A random effects linear regression model with random intercept was applied to test the effects of group, side, and group*side interaction (different covariance structures allowed). Pairwise comparisons were examined and the initial p<.05 level of significance was adjusted with a Bonferroni correction. Non-normally distributed variables were dichotomized but ultimately omitted as the results of logistic regression were unreliable (due to small counts). The relationship between swing time ratio and the timing variables that were significantly different in the asymmetric group was measured by Spearman correlation.

## 3. Results

Six of the 20 and eight of the 29 participants with stroke enrolled in the randomized controlled trial and observational study, respectively, and one control adult were excluded from this analysis due to poor data quality or incomplete collection. Therefore, the final sample size was 35 post-stroke individuals, of whom 27 exhibited TGA, and nine control adults. In total, 485 strides over 188 trials were analyzed.

### 3.1 Demographics and clinical assessment

Participant characteristics are shown in Table 1. Asymmetric individuals generally walked slower and presented with lower CMSA scores compared to those who were symmetric. The inverse relationship of spatiotemporal gait asymmetry with gait speed and motor function/recovery is well-established (Lauzière et al. 2014). There was a higher proportion of primarily left side impairment in the asymmetric than symmetric group, which may be due to the eligibility criteria selecting for individuals without the language or communication deficits that more often accompany right-sided paresis. TGA was in the direction of the more affected side (longer swing time), as is commonly found (Lauzière et al. 2014). Half of the symmetric group presented with longer swing times on the less affected side.

**Table 1.** Demographics and clinical characteristics of participants by group. Values are mean ± standard deviation, median (range), or counts (percentage).

| Variable | Control Adults | Individuals Post-stroke |  |
| --- | --- | --- | --- |
|  |  | Symmetric Group | Asymmetric Group |
| N | 9 | 8 | 27 |
| Age, years | 61.0 ± 8.5 | 62.2 ± 6.0 | 60.7 ± 10.5 |
| Sex |  |  |  |
| Men, n (%) | 3 (33) | 6 (75) | 19 (70) |
| Women, n (%) | 6 (67) | 2 (25) | 8 (30) |
| Time post-stroke, years | - | 2.4 ± 2.2 | 6.0 ± 5.9 |
| More affected side |  |  |  |
| Left, n (%) | - | 3 (38) | 21 (78) |
| Right, n (%) | - | 5 (62) | 6 (22) |
| CMSA<sup>a</sup> |  |  |  |
| Stage of leg | - | 7 (6-7) | 5 (3-7) |
| Stage of foot | - | 6 (5-7) | 4 (1-7) |
| Gait velocity, m/s | 1.06 ± 0.17 | 1.06 ± 0.17 | 0.72 ± 0.20 |
| Swing time ratio | 1.03 ± 0.02 | 1.03 ± 0.02 | 1.39 ± 0.29 |

<sup>a</sup> CMSA: Chedoke McMaster Stroke Assessment.

### 3.2 Temporal gait parameters

Overall, the more affected side of the asymmetric group had the longest swing time (significant interaction effect, p<.0001; Figure 2A). A significant main effect of group was found for absolute stance time (p<.0001; Figure 2A). Since the longer duration of the asymmetric participants could be, at least partly, attributed to gait speed as a confounding factor (Olney and Richards 1996), stance as a percent of stride was also examined. There was a group*side interaction effect (p<.0001; Figure 2B), with the less affected side of the asymmetric individuals having a significantly higher stance proportion (pairwise comparisons p<.0001).

**Figure 2.**
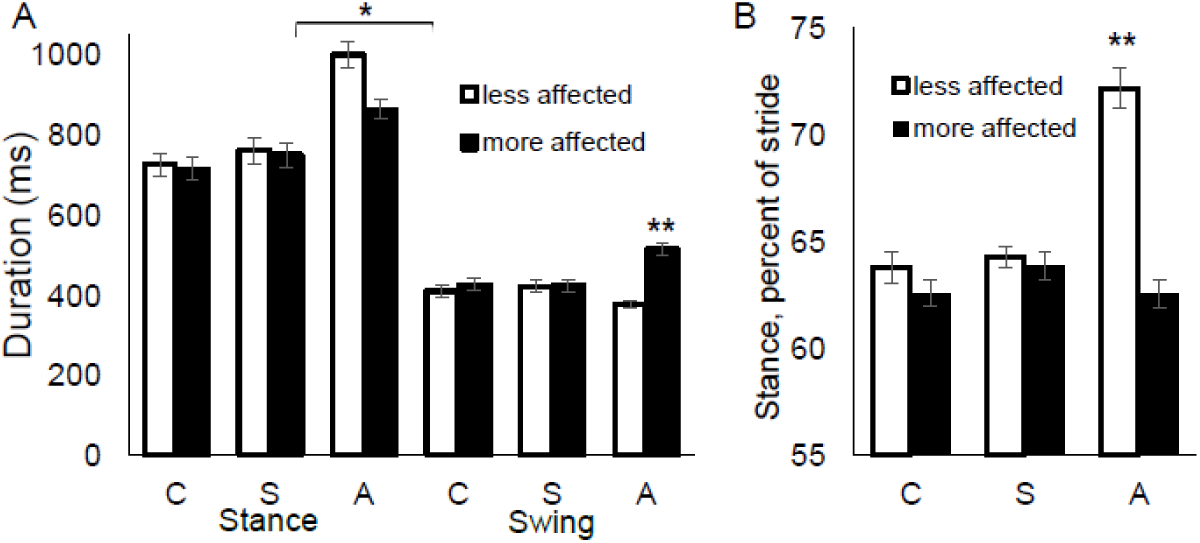
Temporal gait parameters. Stance and swing time (A) and stance percent of stride (B) are shown for each group (C = control adults, S = symmetric, A = asymmetric) by less affected (unfilled bars) and more affected (filled bars) side. The single and double asterisks represent statistically significant between-group difference and group*side interaction effect, respectively (p<.001).

### 3.3 Muscle burst activity

Using the control adult data as a normative template, the following muscle bursts were defined based on the temporal location in the gait cycle (ST=stance, STW=stance-swing transition period, SW=swing phase): TA-ST, TA-STW, TA-SW (Figure 3); MG-ST, MG-SW (occasionally present) (Figure 4); RF-ST, RF-STW (frequently absent), RF-SW (Figure 5); BF-ST1 (early first burst), BF-ST2 (sometimes a later second burst), BF-SW (Figure 6). The prevalence of each burst by group and side is shown in Table 2.

**Table 2.** Timing (percent of stride) and prevalence (n/N) ^a^ of muscle bursts throughout the gait cycle. Values are mean ± standard error % of stride (n/N). n = number of participants exhibiting the burst in ≥50% of strides, N = group sample size (smaller for some sides/muscles if EMG recording was not obtained due to technical issues).

| Burst timing variable | Control Adults |  | Individuals Post-stroke |  |  |  | p-values:<br>group; side;<br>group*side <sup>b</sup> |
| --- | --- | --- | --- | --- | --- | --- | --- |
|  | 'Less affected' <sup>a</sup> | 'More affected' <sup>a</sup> | Symmetric |  | Asymmetric |  |  |
|  |  |  | Less affected | More affected | Less affected | More affected |  |
| Stance (ST) Phase |  |  |  |  |  |  |  |
| TA-ST OFF | 14.7±2.0<br>(9/9) | 15.7±2.6<br>(8/8) | 19.9±3.1<br>(8/8) | 15.4±3.5<br>(8/8) | 32.1±2.6<br>(27/27) | 24.1±2.3<br>(23/26) | **<.0001**; .14; .18 |
| MG-ST ON | 16.6±1.8 | 12.7±2.4 | 18.4±2.5 | 14.8±3.0 | 15.1±2.0 | 7.2±1.7 | .060; **.011**; .49 |
| MG-ST OFF | 50.1±1.2<br>(9/9) | 49.9±1.3<br>(9/9) | 50.6±1.2<br>(8/8) | 47.9±1.5<br>(8/8) | 56.1±2.0<br>(27/27) | 41.0±2.4<br>(25/25) | **.0006** |
| RF-ST OFF | 27.4±2.8<br>(8/8) | 28.5±4.2<br>(9/9) | 27.5±3.7<br>(8/8) | 28.7±3.4<br>(8/8) | 36.0±2.2<br>(27/27) | 29.5±2.1<br>(26/27) | .13; .56; .30 |
| BF-ST1 OFF | 15.5±1.9<br>(9/9) | 16.7±2.0<br>(8/8) | 17.8±2.7<br>(8/8) | 16.9±2.0<br>(8/8) | 19.9±1.3<br>(26/27) | 18.5±1.0<br>(27/27) | .14; .72; .74 |
| BF-ST2 ON | 51.6±5.0 | 49.0±3.2 | 35.5±6.1 | - | 48.4±2.4 | 44.5±2.4 | .45; .36; .83 |
| BF-ST2 OFF | 51.2±2.5<br>(6/9) | 53.7±0.7<br>(4/8) | 42.4±5.0<br>(2/8) | -<br>(0/8) | 60.0±2.1<br>(17/27) | 55.2±2.6<br>(16/27) | .18; .62; .094 |
| Stance-Swing (STW) transition |  |  |  |  |  |  |  |
| TA-STW ON | 59.3±1.2 | 57.8±1.0 | 60.9±0.6 | 57.8±1.3 | 68.4±1.1 | 53.5±1.1 | **<.0001** |
| TA-STW OFF | 72.8±1.2<br>(9/9) | 72.1±1.2<br>(8/8) | 72.3±1.5<br>(7/8) | 75.5±1.4<br>(8/8) | 79.7±1.3<br>(23/27) | 73.0±1.5<br>(23/26) | **.0047** |
| RF-STW ON | 58.0±1.1 | 55.0±0.9 | 55.6±2.4 | 59.3±1.0 | 67.1±1.7 | 56.2±1.6 | **.0021** |
| RF-STW OFF | 66.2±1.6<br>(4/8) | 67.5±2.3<br>(2/9) | 67.4±1.7<br>(4/8) | 66.6±1.5<br>(6/8) | 76.2±1.7<br>(13/27) | 66.2±1.3<br>(12/27) | **.032** |
| Swing (SW) Phase |  |  |  |  |  |  |  |
| TA-SW ON | 88.5±1.8<br>(9/9) | 85.2±2.5<br>(8/8) | 88.3±2.4<br>(8/8) | 88.8±1.1<br>(7/8) | 89.0±1.2<br>(22/27) | 85.4±1.7<br>(15/26) | .59; .12; .56 |
| MG-SW ON | 84.9±3.8 | 76.3±1.6 | 77.1±13.7 | 72.2±9.7 | 75.8±3.6 | 74.7±2.4 | .35; .46; .55 |
| MG-SW OFF | 91.4±3.2<br>(4/9) | 83.7±2.9<br>(4/9) | 85.0±10.2<br>(2/8) | 82.3±9.0<br>(3/8) | 90.2±2.5<br>(8/27) | 88.1±2.4<br>(17/25) | .63; .41; .59 |
| RF-SW ON | 91.4±0.6<br>(8/8) | 91.1±0.6<br>(8/9) | 92.3±1.0<br>(7/8) | 92.8±0.6<br>(5/8) | 90.6±1.4<br>(21/27) | 86.1±1.8<br>(20/27) | **.025**; .18; .13 |
| BF-SW ON | 76.6±2.7<br>(9/9) | 78.9±1.9<br>(8/8) | 84.0±1.8<br>(8/8) | 78.1±3.0<br>(8/8) | 83.7±1.4<br>(25/27) | 80.6±1.6<br>(22/27) | .13; .13; .23 |

<sup>a</sup> Control adult limbs were differentiated by swing time (higher = 'more affected', lower = 'less affected').
<sup>b</sup> Only group\*side interaction p-value shown if significant (bolded).

**Figure 3.**
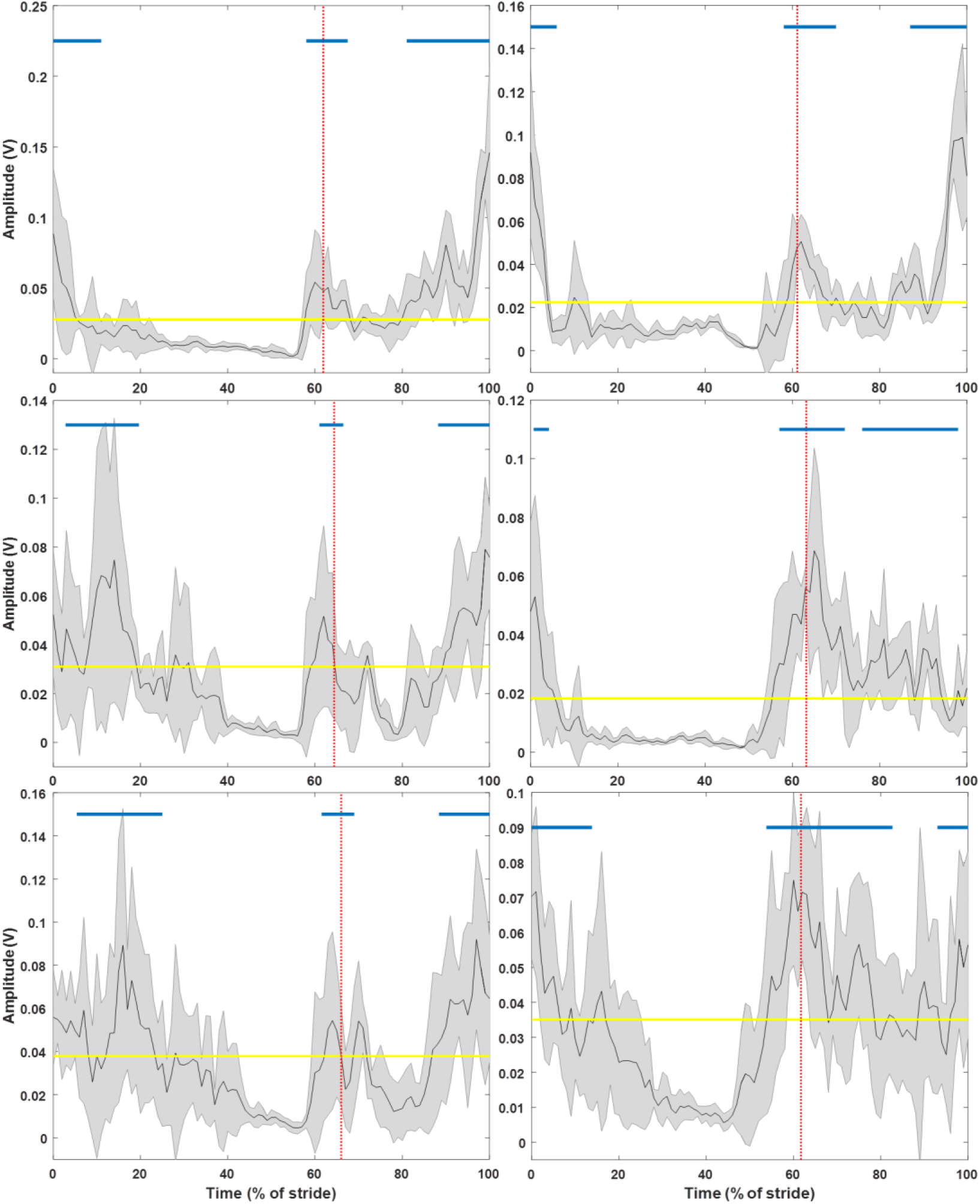
Representative example of tibialis anterior activity from less affected (left panel) and more affected (right panel) side of control adult (top), symmetric individual (middle), and asymmetric participant (bottom). Black line = conditioned EMG signal normalized to the gait cycle and averaged over all strides and trials. Shaded area = standard deviation. Yellow horizontal line = amplitude threshold (trial mean). Red vertical line = average end of stance phase. Blue lines = bursts.

**Figure 4.**
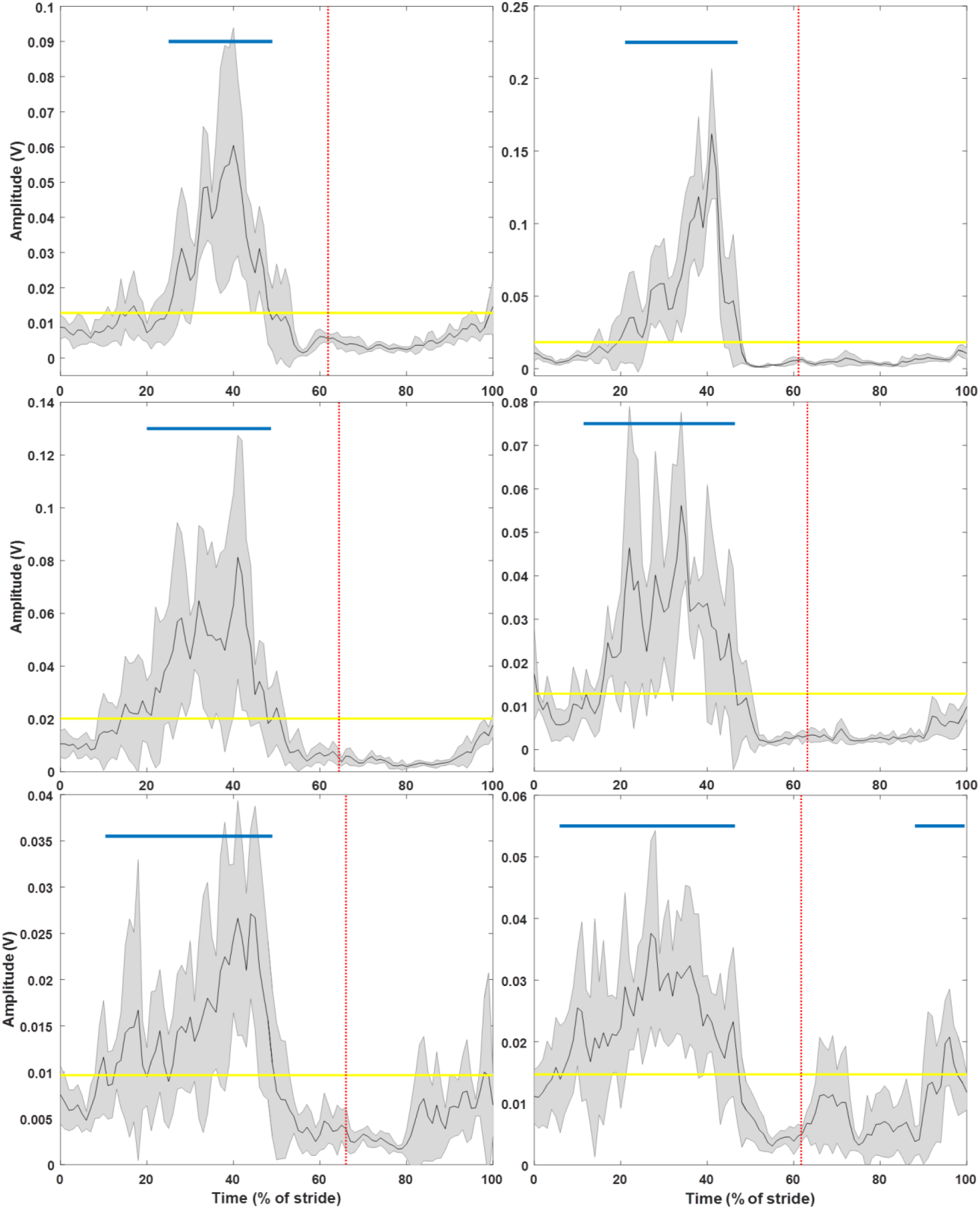
Representative example of medial gastrocnemius activity from less affected (left panel) and more affected (right panel) side of control adult (top), symmetric individual (middle), and asymmetric participant (bottom). Black line = conditioned EMG signal normalized to the gait cycle and averaged over all strides and trials. Shaded area = standard deviation. Yellow horizontal line = amplitude threshold (trial mean). Red vertical line = average end of stance phase. Blue lines = bursts.

**Figure 5.**
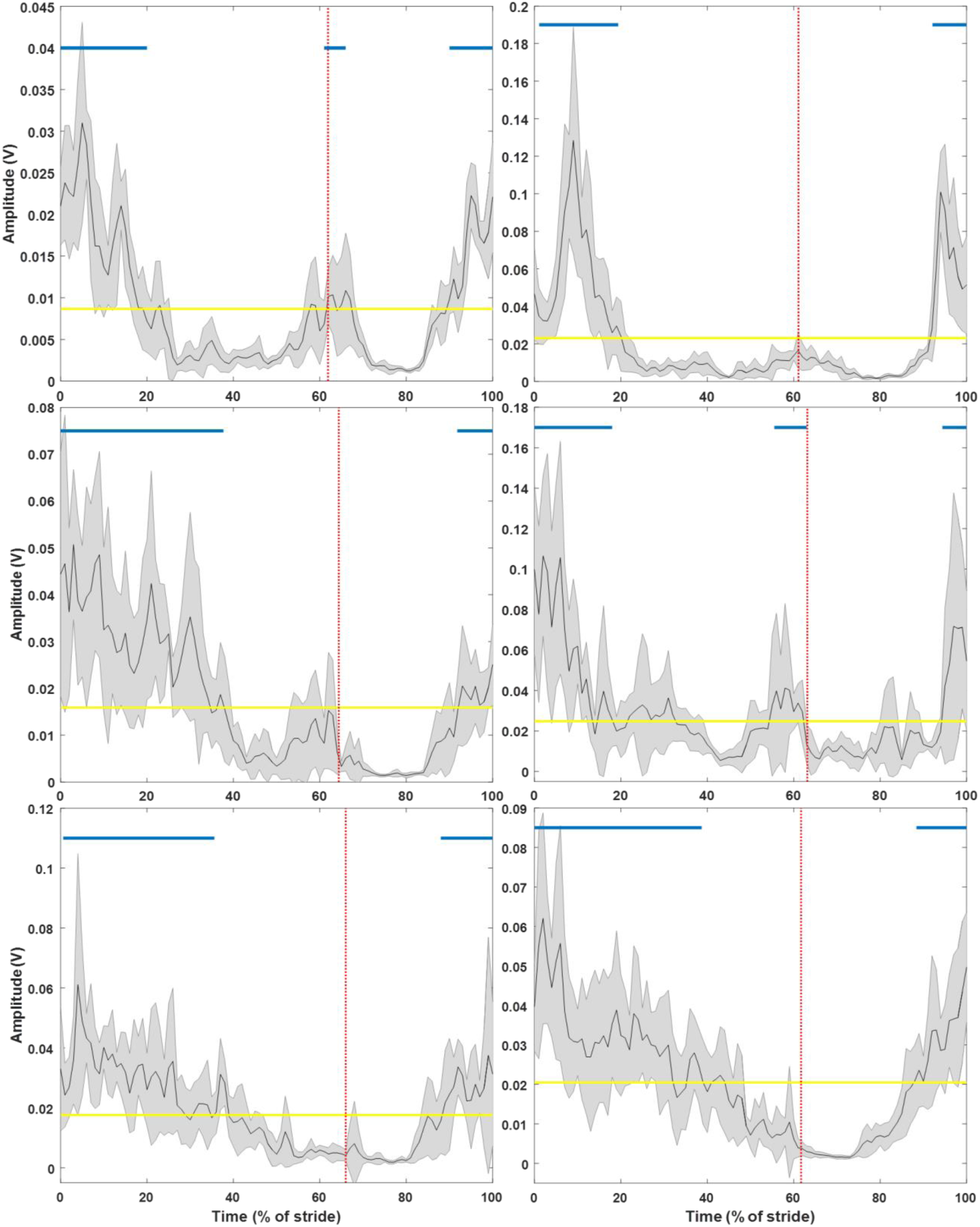
Representative example of rectus femoris activity from less affected (left panel) and more affected (right panel) side of control adult (top), symmetric individual (middle), and asymmetric participant (bottom). Black line = conditioned EMG signal normalized to the gait cycle and averaged over all strides and trials. Shaded area = standard deviation. Yellow horizontal line = amplitude threshold (trial mean). Red vertical line = average end of stance phase. Blue lines = bursts.

**Figure 6.**
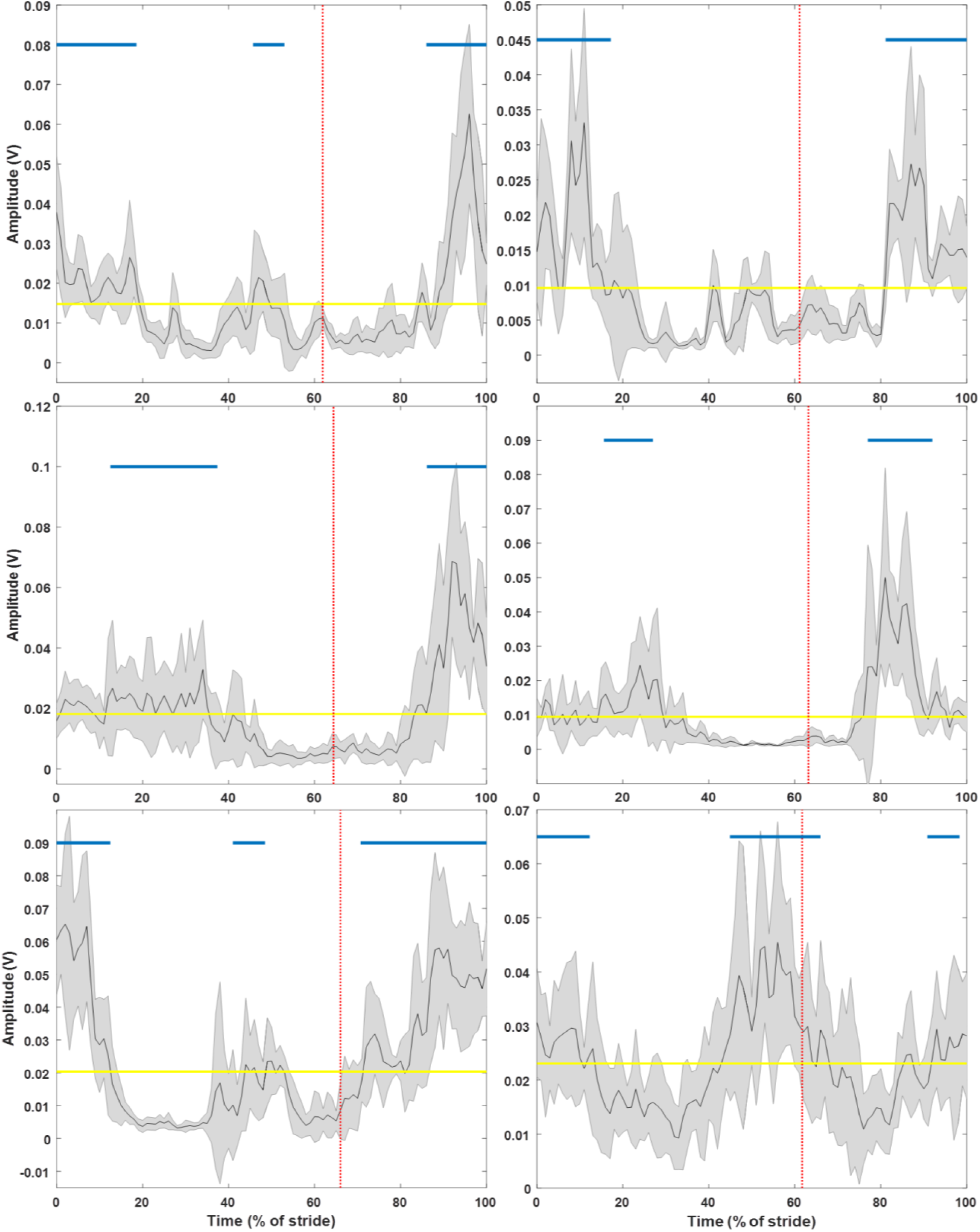
Representative example of biceps femoris activity from less affected (left panel) and more affected (right panel) side of control adult (top), symmetric individual (middle), and asymmetric participant (bottom). Black line = conditioned EMG signal normalized to the gait cycle and averaged over all strides and trials. Shaded area = standard deviation. Yellow horizontal line = amplitude threshold (trial mean). Red vertical line = average end of stance phase. Blue lines = bursts.

The overall muscle activity pattern of the symmetric group was similar to controls, but the later BF-ST2 burst was rarely detected in both lower limbs (significant group effect for the number of BF bursts during stance, p=.039). In the asymmetric group, most participants exhibited a MG-SW burst on the more affected side. A main effect of group (p=.0009) for the number of TA-SW bursts was found; the asymmetric individuals had significantly fewer than the symmetric group (p=.014) and control adults (p=.001). There was also a group effect on the MG-ST burst variable (p=.032) but no significant pairwise differences were found.

### 3.4 Muscle burst timing

ON and OFF timing and the number of participants that exhibited each muscle burst is shown in Table 2. The overall temporal pattern of muscle activity for each group by side combination is visually represented in Figure 7. There was a significant effect for the OFF timing of TA during stance (TA-ST OFF) with the asymmetric group later than the symmetric (p=.0035) and control adults (<.0001). The start of MG activity was significantly earlier in the stance phase (MG-ST ON) on the more affected than less affected side in all groups (p=.011). A significant interaction effect for the end of the MG-ST burst (MG-ST OFF, p=.0006) was found: the more affected side of the asymmetric individuals was early compared to the symmetric and control adults. The stance-to-swing transition burst of TA on the less affected side was shifted significantly later for the asymmetric participants relative to the other groups (pairwise comparisons: TA-STW ON, p≤.0001; TA-STW OFF, p<.01). There was also a significant interaction effect for the timing of the RF-STW burst; when present, it was delayed on the less affected side of the asymmetric individuals (pairwise comparisons: RF-STW ON, p=.0018-.045; RF-STW OFF, p=.037-.29). The start of RF swing phase activity (RF-SW ON) in the asymmetric participants was significantly earlier than the symmetric (p=.020).

**Figure 7.**
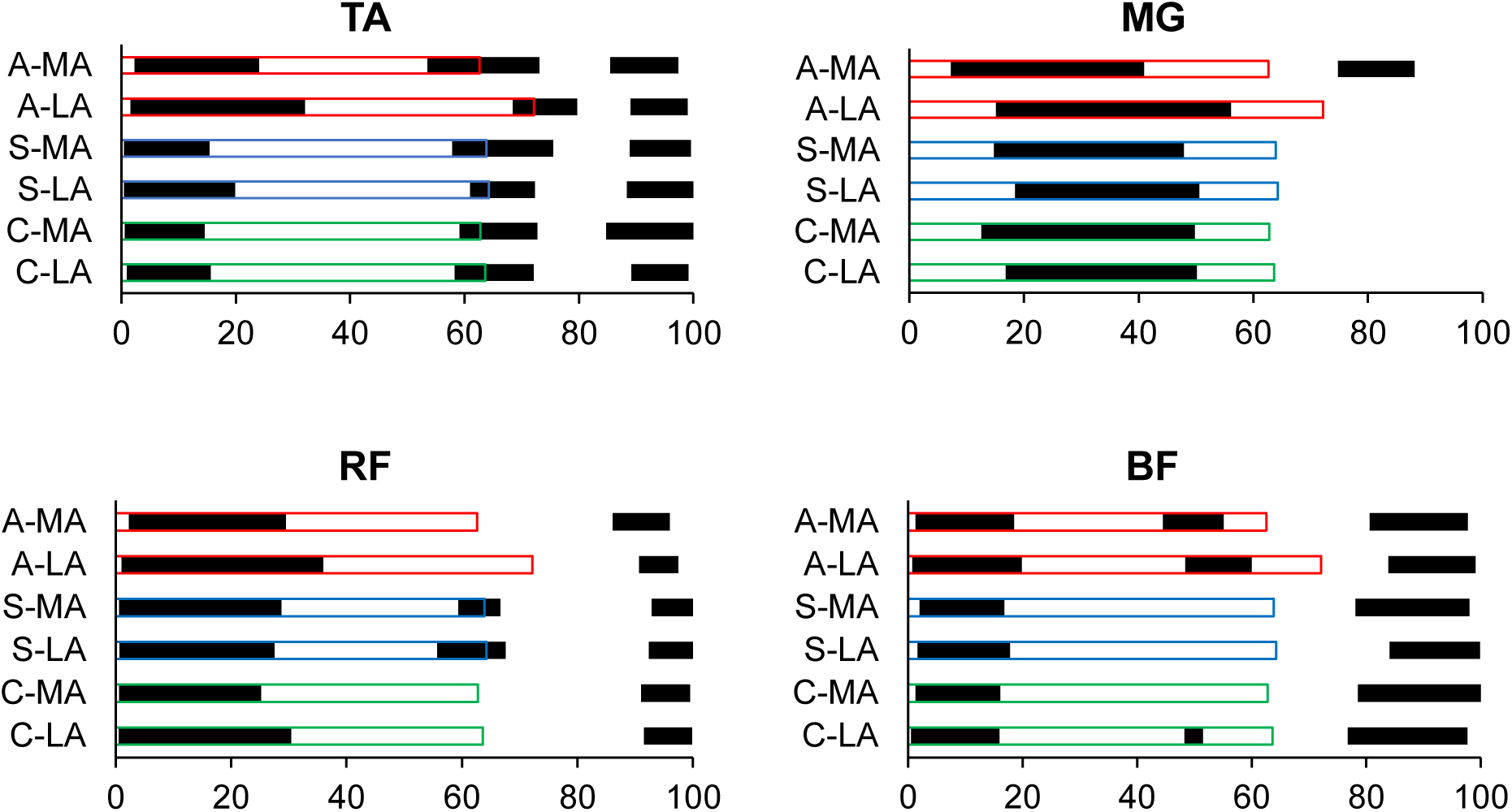
Muscle activity patterns on the less affected and more affected sides of control adults (C-LA and C-MA), symmetric (S-LA and S-MA) and asymmetric (A-LA and A-MA) individuals. Filled bars represent bursts (ON/OFF timing is mean percent of stride) and unfilled bars indicate stance phase.

### 3.5 Correlation between asymmetry and muscle burst timing

Within the asymmetric group, the variables that correlated most with swing time ratio were the stance phase MG burst OFF time on the less affected side (MG-ST OFF: r_s_=0.68, p=<.0001), and the timing of the TA and RF bursts on the less affected side during the stance-to-swing transition (TA-STW ON: r_s_=0.90, p<.0001; TA-STW OFF: r_s_=0.73, p=.0003; RF-STW ON: r_s_=0.86, p=.0001; RF-STW OFF: r_s_=0.87, p=.0001).

## 4. Discussion

The purpose of this study was to explore the neuromuscular mechanisms underlying TGA, which had not yet been directly characterized at this dynamic physiological level. Lower limb EMG during overground walking revealed differences in burst prevalence and on/offset time between persons with TGA, symmetric post-stroke individuals and control adults. Notably, the asymmetric group exhibited fewer TA-SW bursts, early MG-ST OFF timing on the more affected side, and delayed activity of the less affected TA from stance to swing. The findings suggest that certain electromyographic patterns are associated with swing time asymmetry and, therefore, may be important targets for intervention to improve gait quality after stroke.

As a cerebrovascular accident can disrupt descending neural inputs that direct movement, the presence of key muscle activations throughout the gait cycle was determined. The symmetric group exhibited a very low prevalence of the second BF stance burst (BF-ST2), which cannot be explained by slowness associated with reduced hamstrings activity and terminal hip extension (Neptune et al. 2008) because walking speed was equivalent to controls. Late BF activation was only present in two individuals, who also lacked a RF-STW burst, suggesting reciprocal coordination of these knee muscles. In contrast to the other groups, most of the asymmetric participants had an MG burst during the swing phase in the limb with greater impairment. A possible explanation for this excess activity is plantarflexor spasticity, previously found to be associated with TGA (Hsu et al. 2003). The lower number of TA-SW bursts in the group with asymmetry, particularly on the more affected side, may underlie the dorsiflexor weakness reported in earlier work (Lin et al. 2006). Thus, specific gross EMG patterns seem to reflect symmetry status in persons with stroke.

While quantifying muscle bursts provides some useful information about subsequent biomechanical output, timing of neuromuscular activity more directly relates to the resultant parameters associated with TGA. The differences in stance time described above (significant main effects of group and side) are consistent with the delayed asymmetric TA-ST OFF and late MG-ST ON in the less affected limb. Prolonged stance phase activation of TA in the asymmetric participants contributed to considerably greater overlap with the MG burst (17% of stride vs 0-2% in the symmetric and control adults). This period of co-contraction may provide ankle stabilization during the loading response and transition to swing. Earlier MG-ST ON time on the more versus less affected side in both stroke groups suggests a Type I pattern of premature activity, first described by Knutsson and Richards (Knutsson and Richards 1979), that may be caused by calf muscle lengthening as dorsiflexion increases. However, this interlimb difference was also seen in the control adults, which argues against an effect of pathological spasticity, but it does help explain the overall lower stance time on the more affected sides. While MG-ST OFF timing was significantly early in the more impaired limb of individuals with TGA, on average, number and total duration of the burst(s) was not reduced, indicating an intact ability to activate the muscle. In fact, the less affected side of the asymmetric participants had the most and longest-lasting MG-ST bursts - perhaps a compensation for generating enough propulsion to advance through swing phase quickly and reduce the time in single-support on the opposite limb. The late shift of the MG-ST and TA-STW bursts in the asymmetric group’s less affected side likely underlies the greater stance proportion of stride as a major driver of TGA, with the ON/OFF timing highly correlated to swing time ratio. A significant interaction effect and strong correlation with asymmetry was also found for the less affected RF-STW ON and OFF time, but since this burst was present in fewer than half of the asymmetric individuals (potentially related to slow walking (Nene et al. 1999)), it probably has limited impact on gait quality.

Since TGA is a problem of timing, an on/off detection method was used to analyze temporal features of the EMG data, which precluded quantitative interpretation of signal intensity; however, the relationship between swing asymmetry and force-generating capacity has previously been investigated (Kim and Eng 2003). Normalizing to the mean amplitude across a trial has been recommended for reliable task-specific examination, especially in patients with neurological disorders who have difficulty with voluntary contraction (Burden 2010). As the threshold for muscular activation, this may have reduced inter-subject variability, but between-group differences were still found and normative EMG profiles herein generally matched overall reference patterns from the literature (Shiavi 1985; Benedetti et al. 2012). Although only four muscle pairs were measured, taken together, both flexion and extension actions across all three major lower limb joints (i.e., ankle, knee and hip) were represented, so a relatively comprehensive picture of locomotor control could be obtained.

The findings of this study add important mechanistic insight about TGA and have implications for therapeutic management. Previous work has focused on a shortened non-paretic swing phase and proposed underlying impairments of forward propulsion and/or balance control during paretic limb support (Lauzière et al. 2014). For the asymmetric participants herein, stance time was increased overall and as a percentage of their less affected stride, implying a preference for stability through weight-bearing on the ‘better’ leg. Correlational analyses have revealed that TGA has significant associations with measures of standing and dynamic balance (Hendrickson et al. 2014; Lewek et al. 2014); further investigation should explore whether practising these tasks can improve symmetry. The temporal gait parameters can be well-accounted for by the EMG patterns observed, specifically of TA and MG activity around the stance-to-swing transition. Burst timing of the more affected side was generally similar to the control adults, suggesting that neurophysiological deficits in this respect were minimal. While some indirect evidence for the involvement of ankle impairments (weakness, spasticity) was found, swing time ratio was most correlated to the ON/OFF time of muscles on the less affected side, which could be actively generating the asymmetry. Biomechanical compensations in the non-paretic limb are known to develop after stroke and can persist into later stages of recovery (Den Otter et al. 2007; Raja et al. 2012). As a result, it is unclear whether the lower CMSA scores in the group with versus without TGA represent residual neuromotor dysfunction or learned non-use. Muscle activity of asymmetric individuals may have adapted to favour the less affected side for weight-bearing and produce a preferred (consciously or not) gait pattern that was reinforced with ‘practice’.

Asymmetry has been resistant to various interventions, but repetitive task-specific training is a promising way to restore coordination (Hollands et al. 2012). Therapeutic strategies that incorporate temporal cuing for interlimb synchronization may be more effective at promoting favourable adaptation. An early investigation into the effect of rhythmic auditory stimulation demonstrated improvements coinciding with normalization of gastrocnemius activation (Thaut et al. 1993). Another study showed an immediate reduction in asymmetry and electromyographic changes through treadmill walking (Harris-Love et al. 2004), implying an inherent capacity for better use of the more affected limb. Yet other work found invariant timing of paretic muscle activity during rehabilitation, suggesting that functional recovery is due to compensation (Den Otter et al. 2006; Buurke et al. 2008). However, if clinical measures (e.g. walking speed, mobility index, ambulation category) are improved at the expense of symmetry, proactive remediation of coordination patterns should also be a focus to avoid negative consequences for the patient. Ultimately, an individualized approach may be necessary to address the variety of problems that arise from stroke. Incorporating EMG recordings in future research would allow clinical assessment and treatment to directly target the neural input that determines the quality of gait.

## 5. Conclusion

Despite the high prevalence among persons with stroke, gait asymmetry is not well-understood and thus, difficult to manage clinically. To investigate the level of neuromuscular input, EMG from asymmetric, symmetric and control participants while overground walking was directly analyzed. Through this novel and detailed characterization of underlying lower limb muscle activity, important patterns associated with TGA were revealed. Differences in the temporal organization of bursts between study groups may account for specific impairments and compensations in the more and less affected limbs, respectively, that contribute to poor gait quality. The results of this study can be used to inform rehabilitation practice with the goal of promoting symmetry and avoiding negative outcomes after stroke.

## Data Availability

Data are not available; participants did not provide consent for sharing data outside the institution.

## Acknowledgements

Jennifer Wong and Anthony Aqui for help with data collection; Dr. Rahim Moineddin for his technical assistance with statistical analyses.

## Notes

**Funding** This work was supported by a Collaborative Health Research Projects Grant from the Canadian Institutes of Health Research and the National Science Engineering Research Council the Heart and Stroke Foundation [#337523]; Canadian Institutes of Health Research [MOP 133577]. The authors also acknowledge the support of the Toronto Rehabilitation Institute; equipment and space have been funded with grants from the Canada Foundation for Innovation, Ontario Innovation Trust, and Ministry of Research and Innovation. AM holds a New Investigator Award from the Canadian Institutes of Health Research (MSH 141983) and KP holds a salary award from the Heart and Stroke Foundation. Sponsors were not involved in study design and execution or article preparation.

### Competing Interest Statement

The authors have declared no competing interest.

### Funding Statement

This work was supported by a Collaborative Health Research Projects Grant from the Canadian Institutes of Health Research and the National Science Engineering Research Council the Heart and Stroke Foundation [#337523]; Canadian Institutes of Health Research [MOP 133577]. The authors also acknowledge the support of the Toronto Rehabilitation Institute; equipment and space have been funded with grants from the Canada Foundation for Innovation, Ontario Innovation Trust, and Ministry of Research and Innovation. AM holds a New Investigator Award from the Canadian Institutes of Health Research (MSH 141983) and KP holds a salary award from the Heart and Stroke Foundation. Sponsors were not involved in study design and execution or article preparation.

